# Serious Illness Conversations in Older Patients at High Risk of Mortality in Primary Care During the COVID-19 Pandemic: A Quasi-Experimental Study

**DOI:** 10.64898/2026.07.12.26357462

**Authors:** Gabrielle Chicoine, Nathalie Germain, Stéphane Turcotte, Émilie Côté, Véronique Gélinas, France Légaré, Jean-Sébastien Paquette, Annette M. Totten, Michèle Morin, Sharon E. Straus, Patrick Archambault

**Affiliations:** Knowledge Translation Program, Li Ka Shing Knowledge Institute, St. Michael’s Hospital, Unity Health Toronto, Toronto, ON, Canada; Centre de recherche du CISSS de Chaudière-Appalaches, Santé Québec Chaudière-Appalaches, Lévis, QC, Canada; VITAM-Centre de recherche en santé durable, Santé Québec Capitale-Nationale-Universitaire, Université Laval, Quebec City, QC, Canada; Department of Family and Emergency Medicine, Université Laval, Québec, QC, Canada; Santé Québec Lanaudière, Saint-Charles-Borromée, QC, Canada; Department of Family and Emergency Medicine, Université Laval, Quebec City, QC, Canada; Department of Medical Informatics and Clinical Epidemiology, School of Medicine, Oregon Health and Science University, Portland, OR, USA; Department of Anesthesiology and Intensive Care, Division of Critical Care Medicine, Université Laval, Québec, QC, Canada

**Keywords:** advance care planning, end-of-life conversations, serious illness, pandemic, knowledge translation, implementation science, primary care

## Abstract

**Purpose:** Serious Illness Conversations (SICs) are essential to delivering person-centered care for older adults with chronic conditions, but are rarely integrated into routine primary care. To address this gap, we compared the effectiveness of a structured training strategy versus passive dissemination of educational materials on SIC documentation rates during the COVID-19 pandemic.

**Methods:** A quasi-experimental study across 13 primary care clinics in Quebec, Canada. Five clinics received structured team-based Serious Illness Care Program training (intervention group) with a provincially disseminated SIC toolkit and eight received the toolkit only (control group). The primary outcome was the proportion of patients with a documented SIC across three time periods (Period 1, pre pandemic; Period 2, pandemic initial wave; and Period 3, post dissemination of SIC toolkit). We used generalized estimating equations (GEE).

**Results:** Across 13 clinics, 2,368 eligible patients (mean age 75.8 years (*SD* = 7.5), 54% female, with a mean Charlson Comorbidity Index of 4.88 (*SD* = 2)) accounted for 19,134 clinical visits, 49.5% in person and 49.6% virtually. SIC documentation rates were 3.3% (control) and 3.4% (intervention) in Period 1, 9.3% and 4.3% in Period 2, and 6.4% and 4.8% in Period 3, respectively. There was no statistically significant improvement to SIC documentation in the intervention group at Period 2 nor Period 3.

**Conclusion:** Structured training was not more effective than passive dissemination for SIC documentation. Educational interventions must be supported by structural changes, workflow integration, and organizational leadership. Multi-level implementation strategies are needed to embed SICs sustainably into primary care.

## Introduction

Chronic diseases are the leading cause of death worldwide.^1^ High-quality care for older adults with serious chronic illness prioritizes a holistic, person-centered approach focused on quality of life.^2^ Serious Illness Conversations (SIC) about future healthcare decisions should be proactively initiated and frequently reviewed by care teams, in advance of anticipated impairment in the patient decision-making capacity.^3–6^ These proactive and collaborative SICs between patients, family members, and healthcare professionals (HPs) facilitates Advance Care Planning (ACP).^7^

ACP is a communication process that supports patients in understanding and sharing their personal values, life goals, and preferences regarding future medical care, including advance directives.^8,9^ The goal of ACP is to build prognostic awareness while exploring patient hopes and fears to align medical care with their unique priorities during serious and chronic illness.^3^ ACP allows for the revision of patient preferences and related care decisions and reduces unwanted aggressive or life-sustaining interventions at the end of life, hospitalization rates, and healthcare costs.^10–12^ Benefits of ACP discussions include reducing psychological distress in patients and their entourage and improvement in wellbeing, satisfaction with healthcare, and quality of life.^13–17^ For HPs, ACP reduces distress and burden,^18^ partly by addressing uncertainty around the dying trajectory and timing of death.^19,20^

Integration of SIC and ACP into routine clinical practice remains a significant challenge for many HPs and organizations.^21–23^ Most older patients would like to discuss their end-of-life preferences, but only a minority report the opportunity for such conversations with a HP.^24^ This emphasizes an urgent need to be proactive in initiating SICs and ACP. Several barriers for primary care HPs to engage in SIC and ACP exist,^25–28^ including time constraints,^19,20,29^ lack of communication skills and discomfort in dealing with death,^30,31^ lack of adequate training and administrative support,^32,33^ challenges in prognostication and patient identification,^34–36^ conflicting opinions among care team members on who should be responsible for initiating and leading SICs,^37–40^ and the absence of standardized documentation.^41,42^

Primary care is well suited for early and ongoing SICs with patients with chronic illness, as most of their care occurs in this setting.^43–45^ However, SICs are not consistently integrated into primary care workflows and many older patients are not engaged in conversations early enough to make a meaningful impact.^46–49^ Some primary care physicians often delay SICs until late in the disease course,^23,50^ with most SICs occurring approximately one month before death, although most older patients wish to engage earlier.^51^

During the COVID-19 pandemic, the need for early SICs and ACP in primary care clinics and nursing homes became evident as many patients experienced rapid clinical deterioration.^20,23^ In this context, primary care clinics needed to rapidly equip healthcare professionals with the skills to identify patients at high risk of mortality and initiate early SICs. Yet little was known about the most effective way to implement these conversations. We investigated whether structured SIC training or a SIC toolkit disseminated passively would increase SIC documentation rates during the COVID-19 pandemic.

## Methods

### Context

This study is nested within a cluster randomized controlled trial funded by the Patient-Centered Outcome Research Institute (PCORI®: PLC-1609-36277, NCT03577002).^18,45,52,53^ The trial aimed to compare the effectiveness of a team-based vs. clinician-focused approach for implementing the SIC training component in 40 primary care clinics in the USA and Canada, recruited from seven Practice-Based Research Networks (PBRNs) within the Meta-LARC consortium. Both trial interventions were based on the SIC training program, a multicomponent intervention developed by Ariadne Labs (www.ariadnelabs.org) to promote systematic SIC implementation. Clinics were trained from May to October 2019 before the COVID-19 pandemic under the supervision of Oregon Health & Science University (OHSU) and Université Laval.

In response to the pandemic, the Quebec Ministry of Health released a basic SIC toolkit, developed by a committee of researchers and knowledge users, to provide clinical care guidelines for initiating SICs.^54^ The toolkit was disseminated via a passive strategy, posted on the Ministry’s website and shared through a mass email campaign signed by the Deputy Health Minister. No other implementation strategy was employed.

We hypothesized that primary care clinics in which HPs had received either one of the structured SIC trainings (team-based or clinician-focused) during the Meta-LARC trial would be more successful in increasing SIC documentation rates during the COVID-19 pandemic than clinics who only received the SIC toolkit passively.

### Study Design

We used a quasi-experimental design to compare the impact of SIC training versus passive dissemination on SIC documentation rates during the COVID-19. A medical chart review of older adults with at least one chronic illness was conducted from September 2019 to October 2020. We followed the Transparent Reporting of Evaluations with Nonrandomized Designs (TREND) Statement (Supplemental Table 1).^55^ Ethics approval was granted by the Research Ethics Committee at Centre intégré de santé et de services sociaux de Chaudière-Appalaches, Quebec (Project # MP-23-2021-800).

We used a two-phase sampling approach. Six clinics trained in SIC during the Meta-LARC trial were invited; five participated as intervention sites. From 37 eligible control clinics, eight were selected using stratified sampling based on university affiliation, patient volume, and the 2016 Material and Social Deprivation Index (MSDI) of their catchment area.^56^ Ministry data from 2019 and clinic postal codes were used to determine patient volume and MSDI, respectively.

Control clinics from three university networks were selected to match intervention sites on volume and deprivation. Records from all 13 clinics were then randomly sampled to ensure representation by socioeconomic status and clinic size. Clinics where HPs received the Meta-LARC study SIC training were the intervention group. Clinics who only passively received the Quebec SIC toolkit without any additional SIC training were the control group.

Community-dwelling adults (≥65 years) who had at least one visit with a HP in the 13 participating clinics between September 2019 and October 2020 were eligible. Visits could be in person or virtual (via phone or online platform). Participants had at least one chronic condition identified from COVID-19 vulnerability criteria based on the International Severe Acute Respiratory and Emerging Infections Consortium cohort study.^57^ Adults in retirement homes, nursing homes, or long-term care homes were also included.

The primary outcome was the proportion of eligible patients with documented SICs during clinic visits, assessed in three pre-defined periods. Period 1 included the six months preceding the onset of the COVID-19 pandemic (September 12, 2019 to March 12, 2020). Period 2 extended from the start of the pandemic until the release and province-wide email distribution of the basic SIC toolkit by the Quebec Ministry of Health (March 13, 2020 to April 24, 2020). Period 3 included the six months following dissemination of the SIC toolkit (April 25, 2020 to October 25, 2020).

SIC documentation was determined based on Sinuff’s framework,^58^ including 34 indicators for evaluating end-of-life communication. Standardized SIC documentation (e.g., care plans, advance directives) or any unstructured ACP note indicating a conversation about patient goals and values was considered as documented SIC. Notes could be written by any HP involved in patient care.

Sociodemographic and clinical data, along with clinical visit and clinic characteristics, were collected and stored securely in a REDCap database (Vanderbilt University, TN, USA).^59^ Medical residents and research staff, not blinded to group allocation, extracted data from electronic medical records using a structured form and were supervised by an experienced research coordinator.

### Statistical Analyses

Sample size calculations were based on clinic workflow data, estimating that 1,892 medical records were needed to detect a 10% difference in SIC documentation rates, with a Bonferroni-corrected alpha of 0.017 and 80% power. The estimated baseline SIC documentation rate in the intervention group was 30%, and the same estimate was used for the control group.

Generalized linear models (GLMs) with generalized estimating equations (GEEs) were used to examine the effect of training on SIC documentation across the three periods. This approach accounts for repeated measures and is robust to mis-specified correlation structures and overdispersion.^60^ Main effects of group, time, and the group × time interaction were assessed. The initial model was adjusted for covariates (age, sex, residence, social and material deprivation index, Charlson Comorbidity Index,^61^ baseline proportion of documented SICs, in-person versus virtual visits, and whether or not the visit was with a physician) as fixed between-participant effects. Tukey-Kramer adjustments were used for pairwise comparisons of group and time interactions. Odds ratios with 95% confidence intervals (CIs) were calculated.

Descriptive statistics were used for sociodemographic and clinical data, and characteristics of primary care practices. Continuous variables were presented as means and standard deviations; categorical variables as frequencies. All analyses were conducted using SAS software, version 9.4 (SAS Institute, Inc., NC, USA), by an experienced biostatistician.

## Results

From 13 clinics, 3,740 medical records were randomly selected, and 2,368 patients met inclusion criteria (see Supplemental Figure 1 for study flow diagram). These patients had 19,134 clinical visits (9474 (49.5%) in person and 9493 (49.6%) virtually via phone or online) across the three study periods. Most patients were female (54%) with an average age of 75.8 years (SD = 7.5) and a mean Charlson Comorbidity Index of 4.88 (SD = 2.04). Table 1 presents patient demographics and clinical visit characteristics by study group and for the full sample.

**Table 1.**
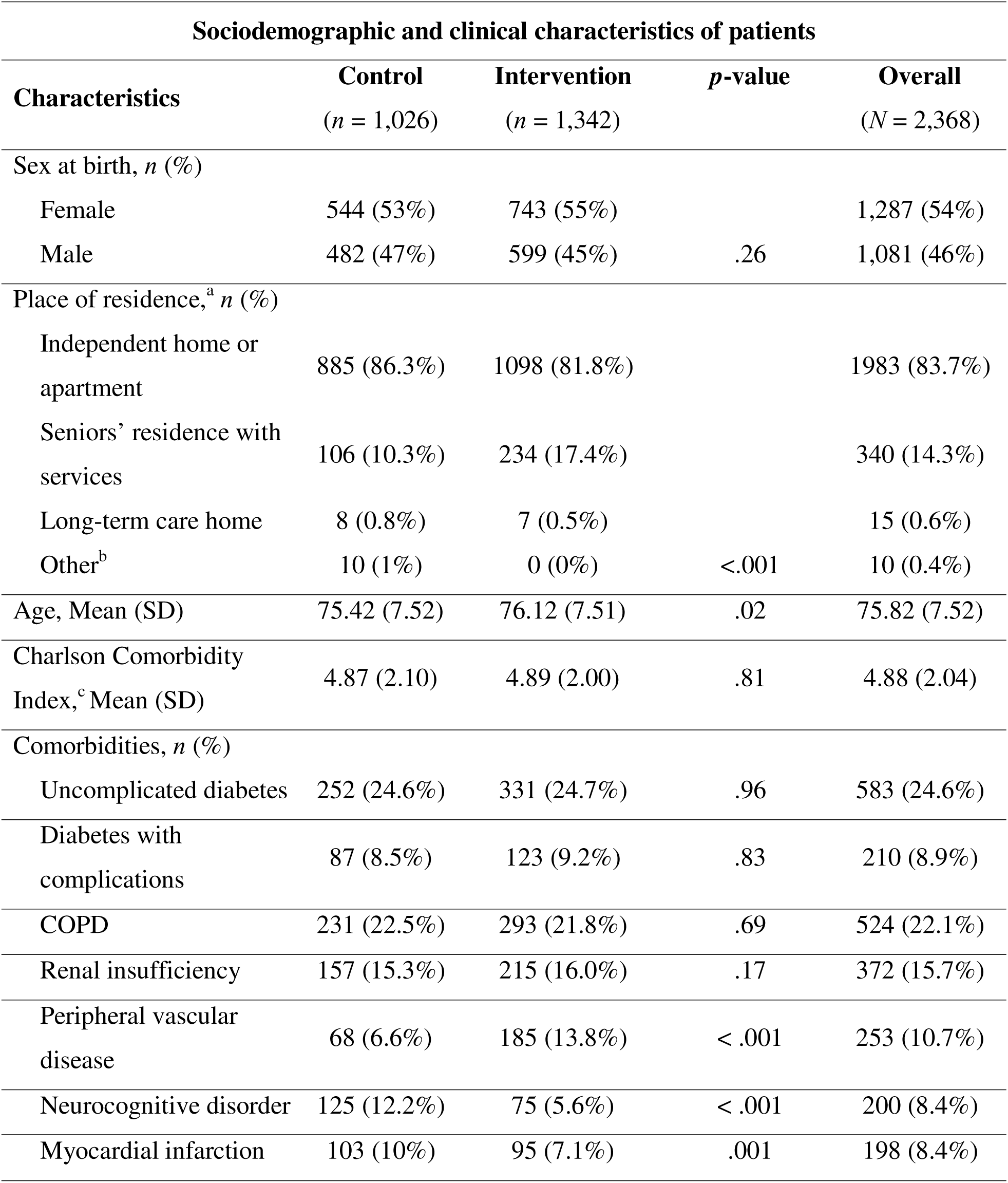
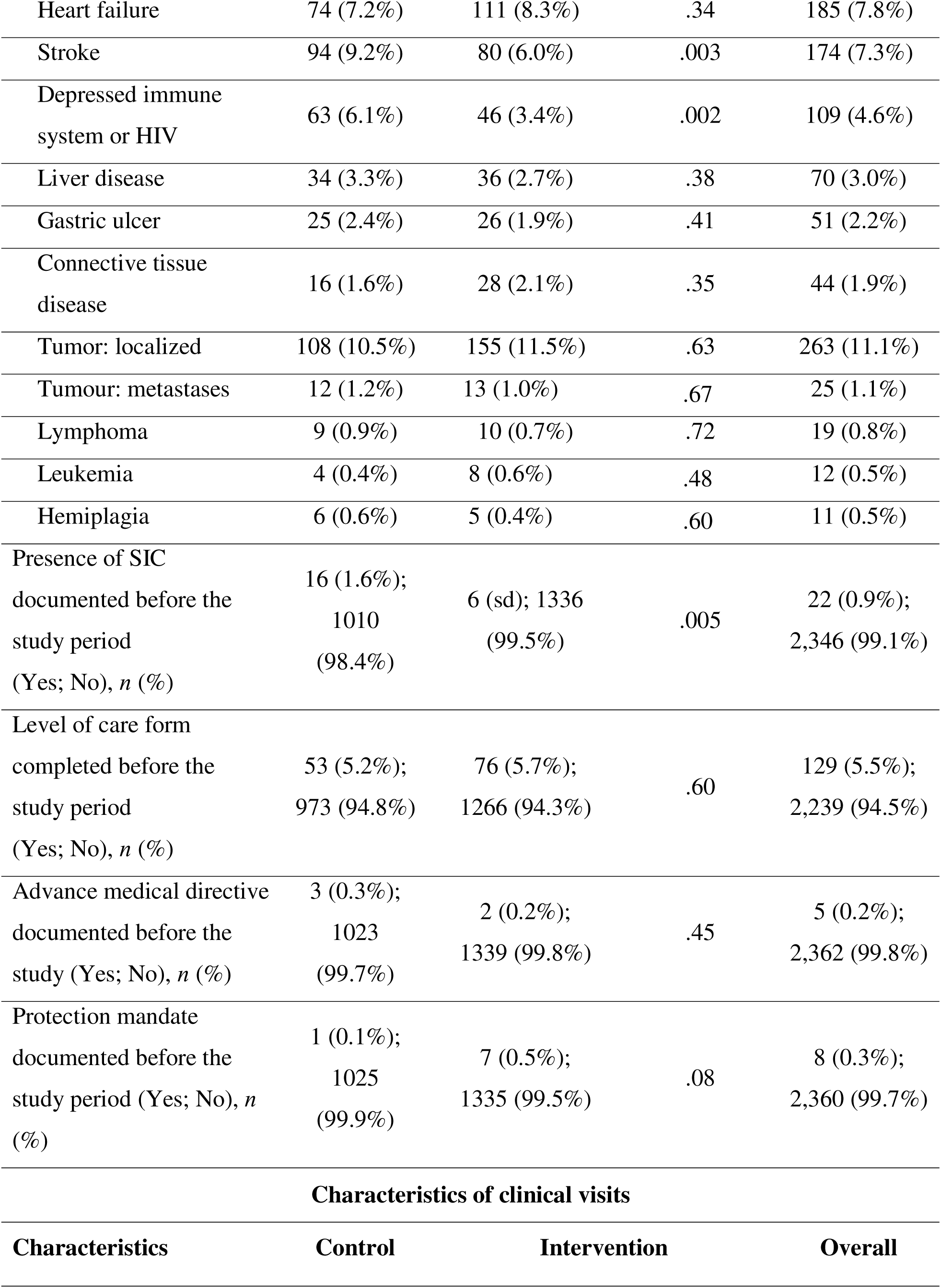
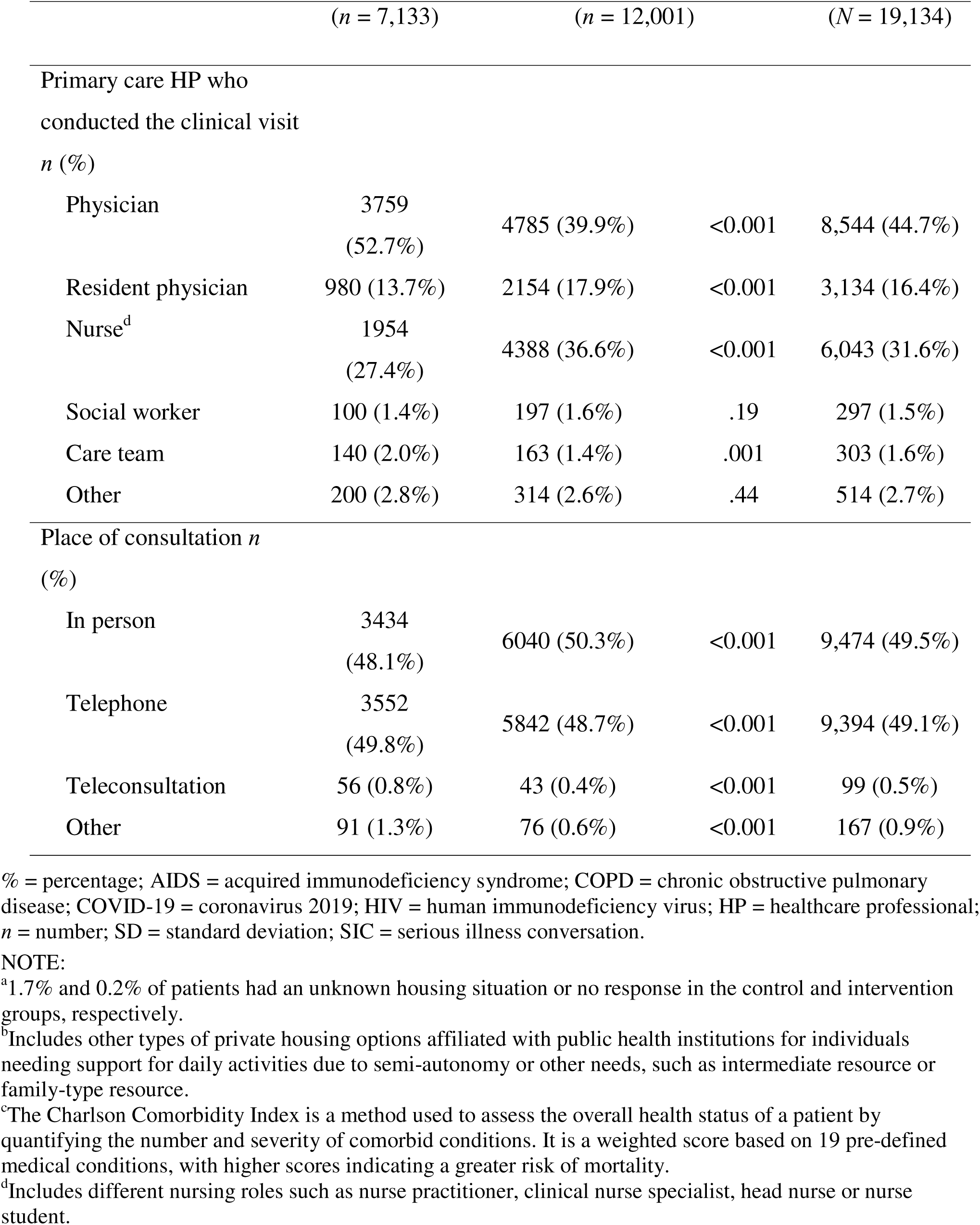
Patient and clinical visit characteristics, by study group and overall.

For both groups, SIC documentation increased over time. In Period 1, 3.3% (*n* = 25/747) of visits in the control group and 3.4% (*n* = 37/1091) in the intervention group had SIC documentation. In Period 2, the control group had 9.3% (*n* = 51/551), and the intervention group had 4.3% (*n* = 33/770). In Period 3, documentation was 6.4% (*n* = 55/865) for the control group and 4.8% (*n* = 55/1138) for the intervention group. GEE model results indicated a significant intervention × time interaction. Compared with Period 1 (Table 2), the intervention group showed no statistically significant change in the adjusted odds of SIC documentation at Period 2 (OR = 1.38, 95% CI 0.70–2.74, *p* = .76) and no significant difference between Period 2 and Period 3 (OR = 1.08, 95% CI 0.54–2.17, *p* = .99). Conversely, the control group had a statistically significant increase in SIC documentation at Period 2 (OR = 3.64, 95% CI 1.81–7.33, *p* < .0001), but no significant difference between Period 2 and Period 3 (OR = 1.08, 95% CI 0.54–2.17, *p* = .99). For all pairwise differences, see Figure 1.

**Figure 1.**
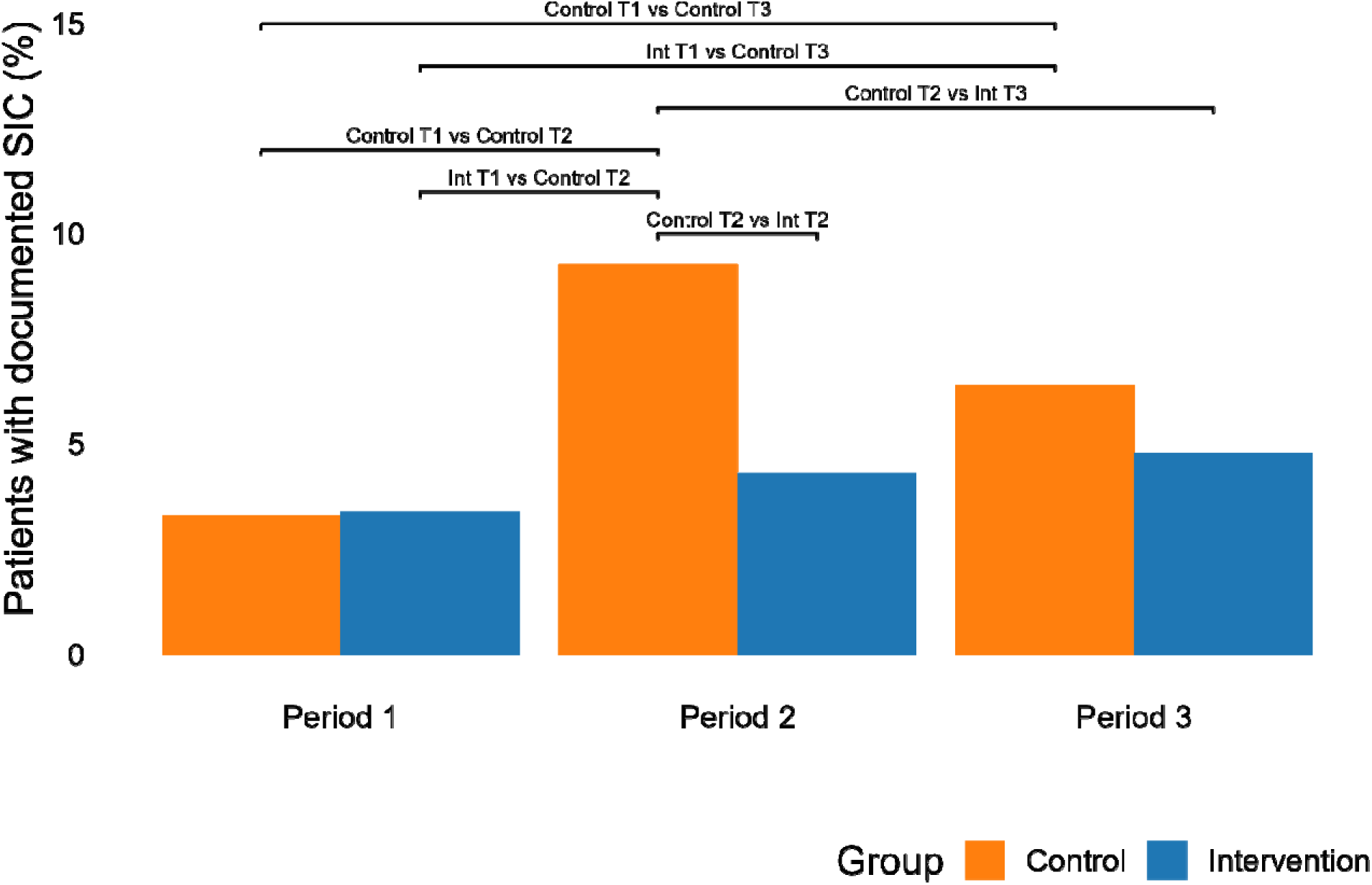
Percentage of patients with documented serious illness conversation (SIC), by intervention group and time period (*N* = 2,368). Horizontal lines between bars denote statistically significant pairwise differences, annotations describe the comparisons.

**Table 2.** Odds ratios (ORs) and 95% confidence intervals (CIs) in for base model and for covariate-adjusted (adjusted model) analysis of SIC documentation by intervention and time.

| Variable | Base model<br>OR (95% Confidence<br>interval) | *p*-value | Adjusted model<br>OR (95% Confidence<br>interval) | *p*-value |
| --- | --- | --- | --- | --- |
| Group<br>Intervention vs control | 0.69 (0.52–0.92) | .01 | 0.67 (0.50–0.90) | .01 |
| Period<br>P2 vs P1<br>P3 vs P1 | 1.87 (1.36–2.58)<br>1.70 (1.24–2.32) | <.0001 | 2.24 (1.49–3.37)<br>1.80 (1.21–2.66) | <.0001 |
| Interaction Group x Period<sup>1</sup><br>Intervention P2 vs P1<br>Intervention P3 vs P2<br>Control P2 vs P1<br>Control P3 vs P2 | 1.21 (0.62–2.33)<br>1.22 (0.62–2.36)<br>2.90 (0.62–2.36)<br>0.68 (0.40–1.13) | 0.02 | 1.38 (0.70–2.74)<br>1.08 (0.54–2.17)<br>3.64 (1.81–7.33)<br>0.60 (0.34–1.04) | < .011 |
| Age |  |  | 1.08 (1.06–1.11) | < .0001 |
| CCI |  |  | 1.20 (1.12–1.28) | < .0001 |
| Visits led by physicians<br>(reference level = visit led by<br>another professional) |  |  | 1.80 (1.26–2.57) | .001 |
| In-person visits<br>(reference level = virtual or phone<br>consultations) |  |  | 1.57 (1.10–2.23) | .012 |
| Socially advantaged site |  |  | 0.73 (0.54–0.99) | .045 |
| (reference level = socially disadvantaged) |  |  |  |  |
| Male sex at birth<br>(reference level = female sex at birth) |  |  | 1.28 (0.95–1.74) | .11 |
| Lives at home<br>Lives in RPA<br>(reference level = Other) |  |  | 0.32 (0.15–0.69)<br>0.40 (0.18-0.90) | .001<br>.022 |

<sup>1</sup>Interaction terms denote the interaction between the effects of group (Control or Intervention) and Time (Period 1 (P1), Period 2 (P2), Period 3 (P3)). Only multiple comparisons of interest were presented to meet the objective.

Significant model covariates included age, Charlson Comorbidity Index, visits led by physicians, in-person visits compared with virtual or telephone consultations, socially advantaged sites, and living situation (See Table 2). Baseline differences in the proportion of SICs performed *before* the study was not statistically significant (OR = 1.75, 95% CI = 0.80–3.85, *p* = .16) and this predictor was removed from the final model. In sensitivity analyses, we removed an outlier control site, which had SIC documentation rates of 0.9%, 12.0%, and 20.3% for Periods 1, 2, and 3, respectively, compared to control group rates of 3.3%, 9.3%, and 6.4%. After removing the outlier control site, results remained consistent for the intervention group both Period 2 (OR = 1.44, 95% CI 0.73–2.86, *p* = .64) and Period 3 (OR = 1.05, 95% CI 0.35–2.12, *p* = .99).

## Discussion

### Principal Findings

We assessed the impact of two implementation strategies on SIC documentation rates among older, high-risk patients in primary care during the COVID-19 pandemic in Quebec. We hypothesized that clinics with prior SIC training would show higher documentation rates compared to those exposed only to passive educational materials. Contrary to this, the intervention group showed lower SIC documentation rates than the control group, suggesting that structured training may not be superior to passive dissemination.

Our findings challenge the assumption that formal training is more effective for SIC adoption. While passive dissemination is generally seen as ineffective, recent research suggests it can work under certain conditions. Key factors for successful passive dissemination include professional confidence, an identified champion, and sufficient resources^62^. Although our study was not designed to verify if these conditions were present in the control sites, these factors may have enabled effective practice change in these sites. Heavy workloads and limited time for ACP in primary care, along with the influence of organizational culture, have been identified as key barriers.^28,63–65^ Strategies that integrate these discussions into time-pressured workflows are more likely to succeedl^21^ and therefore organizational support is a prerequisite for conducting SICs.

Both structured training and passive dissemination resulted in modest improvements in SIC documentation, with the control group showing the most significant gains. This suggests that training or educational materials alone may not drive meaningful behavior change, and that strategies closer to decision-making, such as feedback and reminders, are more effective. In studies on ACP, reminder prompts and electronic medical record tools improved documentation rates, with one study reporting an increase from 0% to 29% using physician prompts^66^ and another from 6% to 56% using an email notification from a machine learning model assessment of the patient record.67 Patients in this study with ACP were more than twice as likely to have code status reduced with ACP documentation.^67^ These findings highlight the importance of combining education with implementation tools like SIC reminders and patient identification tools. Future initiatives should equip healthcare providers with tools that can be integrated into routine practice.

SIC documentation rates were low at baseline in both groups but increased sharply in the control group, while remaining stable in the intervention group. This suggests that barriers at intervention sites may have hindered SIC adoption, while strategies in the control sites, possibly influenced by the pandemic, facilitated adoption. Some of these barriers may be related to environmental, human resource and organizational factors, which include lower socioeconomic status, personnel turnover, time constraints, lack of leadership support, and inadequate infrastructure.^21,28,35,52,64,65,68–70^

Our group^71^ found that barriers in the opportunity domain of the COM-B Model^72^ were particularly influential for influencing intentions to conduct SICs. Consequently, modifications targeting environmental changes could improve the success of future interventions to improve SIC documentation. Our results suggest that educational interventions alone cannot address environmental and resource-related barriers; organizational adjustments may also be necessary. In line with previous research,^71^ we found that physician-led visits in the control group increased the likelihood of documenting SICs. Physicians may have been especially responsive to the ministry’s pandemic call to action due to professional and ethical obligations. Increasing SIC rates may require empowering other healthcare professionals, such as nurses, to lead these discussions.

### Limitations

This study has limitations. First, we used a quasi-experimental design without group randomization, which makes it difficult to show differences in comparative effectiveness, especially when interventions are similar.^55^ While a randomized controlled trial would have been preferable, logistical constraints were too great.

Second, this study compared two implementation strategies at the organizational level, with primary care clinics as the unit of allocation and analysis rather than individual healthcare professionals. The impact of these strategies may depend on their reach within each organization, which could not be measured directly. Although patient demographics were balanced between groups, differences in healthcare professional characteristics could not be assessed and may have influenced the comparison of the primary outcome.

Third, the study was conducted during a period of considerable instability, with evolving work environments, policies, and staffing during the COVID-19 pandemic. These conditions may have influenced SIC documentation and introduce potential historical bias. The extended study period also raises the possibility of contamination between organizations due to staff movement. To address these risks, we incorporated relevant covariates and conducted sensitivity analyses.

Qualitative data could have offered additional contextual insight into variations in the results and factors influencing SIC adoption.

## Conclusion

Advance care planning (ACP) is an evidence-based intervention that helps ensure future care aligns with patient preferences and is particularly amenable to primary care where older adults with serious chronic illness receive most of their care. Serious illness conversations (SICs) support the development of advance care plans reflecting these values. This study compared structured SIC training with passive educational materials to identify strategies to increase SIC documentation during the COVID-19 pandemic in Québec. Although the structured training intervention did not outperform passive education alone, the findings highlight persistent environmental and professional barriers to changing clinician behavior. Strong leadership, organizational support, and broader interprofessional engagement appear essential for embedding SICs into routine practice. Future initiatives should combine skills training with system-level changes to address workflow constraints and support sustainable improvements in patient-centered primary care.

## Data Availability

All data produced in the present study are available upon reasonable request to the authors.

## List of abbreviations

ACP: Advance care planning
CI: Confidence interval
CFPC: College of Family Physicians of Canada
GEE: Generalized estimating equations
GLM: Generalized linear models
HPs: Healthcare professionals
Meta-LARC: Meta-network Learning and Research Center
MRC: Medical Research Council
MSDI: Material and Social Deprivation Index
OR: Odds ratio
PBRNs: Practice-Based Research Networks
PCORI: Patient-Centered Outcomes Research Institute
SD: Standard deviation
SIC: Serious illness conversation
TREND: Transparent Reporting of Evaluations with Nonrandomized Designs

## Acknowledgements

We would like to express our gratitude to all the resident physicians and medicine students for their invaluable assistance in the data collection process, including Philippe Aubert, Alex Bédard, Fabien Bergeret, Vincent Boun, Gabriel Breault, Baptiste Champvillard, Dominique Chabot, Jonathan Cormier, Laurie Couture, Louis-Olivier Cyr, Vincent Dubuc, Michelle Forcier, Fannie Gélinas-Gascon, Camille Gagnon-Potvin, Mathilde Leblond, Catherine Lepage, Jamie-Lee Lépine, Félix Lajoie-Harvey, Mathilde Lamontagne, Flavi Mansour, Camille Morin, Cindy Pelletier-Caron, Alexandre Poitras, Frédérique Préfontaine-Racine, Justin Parent, Olivia Ptito-Taras, Salsabil Selmi, Myriam Sentürk-St-Onge, and Samuel Villeneuve. We also extend our appreciation to our local partners and the dedicated members of our research team whose ongoing commitment and support were essential to the conduct of this study; the following individuals played key roles in the project: Diane Anglehart, Marie Authier, Andréanne Bernatchez, Anne-Pascale Côté, Yves Couturier, Julie Deschambault, Victoria Dorimain, Souleymane Gadio, Raphaëlle Giguère, Sabrina Guay-Bélanger, Johanne Hébert, Seiko Izumi, Lucille Juneau, Hélène Landry, Brigitte Laflamme, Mathew Menear, Annie Miron, Nadjib Mokraoui, Audrey Nolet, Jérôme Patry, Esther Poiré, Sylvie Plourde, Alfred Kodjo Toi and Ivan Ubeda.

## Authors’ contributions

GC: Contributing to data interpretation and analysis, writing of original draft, editing and reviewing of the manuscript.

NG: Contributing to data interpretation and analysis, contributing to writing of original draft, editing and reviewing of the manuscript.

ST: Contributing to conceptualization of the study, leading all data analysis.

EC: Contributing to conceptualization of the study, contributing to coordinating research activities, contributing to editing and reviewing of the manuscript.

VG: Contributing to conceptualization of the study, contributing to coordinating research activities, contributing to editing and reviewing of the manuscript.

FL: Contributing to conceptualization of the study, contributing to editing and reviewing of the manuscript.

JSP: Contributing to conceptualization of the study, contributing to editing and reviewing of the manuscript.

AT: Contributing to conceptualization of the study, contributing to editing and reviewing of the manuscript.

MM: Contributing to conceptualization of the study, contributing to editing and reviewing of the manuscript.

SES: Contributing to conceptualization of the study, contributing to editing and reviewing of the manuscript.

PA: Conceptualization of the study, leading all study procedures, overseeing data collection and management, contributing to data interpretation and analysis, editing and reviewing of the manuscript.

## Availability of data and materials

The datasets used and/or analyzed during the current study are available from the corresponding author on reasonable request.

