## Supplement for "Serious Illness Conversations in Older Patients at High Risk of Mortality in Primary Care During the COVID-19 Pandemic: A Quasi-Experimental Study"

### SUPPLEMENTARY MATERIAL

#### Supplemental Table 1. Transparent Reporting of Evaluations with Nonrandomized Designs (TREND) Statement checklist^70^

| **Paper Section/Topic** | **Item #** | **Descriptor** | Reported? | |
| --- | --- | --- | --- | --- |
|  |  |  |  | Page # |
| **TITLE and ABSTRACT** | | |  |  |
| Title and Abstract | 1 | - Information on how units were allocated to interventions | x | 3 |
|  |  | - Structured abstract recommended | x | 3 |
|  |  | - Information on target population or study sample | x | 3 |
| **INTRODUCTION** | | |  |  |
| Background | 2 | - Scientific background and explanation of rationale | x | 4 |
|  |  | - Theories used in designing behavioral interventions | x | 5 |
| **METHODS** | | |  |  |
| Participants | 3 | - Eligibility criteria for participants, including criteria at different levels in recruitment/sampling plan (e.g., cities, clinics, subjects) | x | 5-8 |
|  |  | - Method of recruitment (e.g., referral, self-selection), including the sampling method if a systematic sampling plan was implemented | x | 5-8 |
|  |  | - Recruitment setting | x | 5-8 |
|  |  | - Settings and locations where the data were collected | x | 5-8 |
| Interventions | 4 | - Details of the interventions intended for each study condition and how and when they were actually administered, specifically including: | x | 5-8 |
|  |  | - Content: what was given? | x | 35 |
|  |  | - Delivery method: how was the content given? | x | 5-8 |
|  |  | - Unit of delivery: how were subjects grouped during delivery? | x | 5-8 |
|  |  | - Deliverer: who delivered the intervention? | x | 5-8 |
|  |  | - Setting: where was the intervention delivered? | x | 5-8 |
|  |  | - Exposure quantity and duration: how many sessions or episodes or events were intended to be delivered? How long were they intended to last? | x | 5-8 |
|  |  | - Time span: how long was it intended to take to deliver the intervention to each unit? | x | 5-8 |
|  |  | - Activities to increase compliance or adherence (e.g., incentives) | x | 5-8 |
| Objectives | 5 | - Specific objectives and hypotheses | x | 6 |
| Outcomes | 6 | - Clearly defined primary and secondary outcome measures | x | 7 |
|  |  | - Methods used to collect data and any methods used to enhance the quality of measurements | x | 7 |
|  |  | - Information on validated instruments such as psychometric and biometric properties | x | 7 |
| Sample size | 7 | - How sample size was determined and, when applicable, explanation of any interim analyses and stopping rules | x | 7-8 |
| Assignment method | 8 | - Unit of assignment (the unit being assigned to study condition, e.g., individual, group, community) | x | 5-7 |
|  |  | - Method used to assign units to study conditions, including details of any restriction (e.g., blocking, stratification, minimization) | x | 5-7 |
|  |  | - Inclusion of aspects employed to help minimize potential bias induced due to non-randomization (e.g., matching) | x | 5-7 |
| Blinding (masking) | 9 | - Whether or not participants, those administering the interventions, and those assessing the outcomes were blinded to study condition assignment; if so, statement regarding how the blinding was accomplished and how it was assessed | x | 7 |
| Unit of Analysis | 10 | - Description of the smallest unit that is being analysed to assess intervention effects (e.g., individual, group, or community) | x | 7 |
|  |  | - If the unit of analysis differs from the unit of assignment, the analytical method used to account for this (e.g., adjusting the standard error estimates by the design effect or using multilevel analysis) | x | 6-7 |
| Statistical methods | 11 | - Statistical methods used to compare study groups for primary methods outcome(s), including complex methods for correlated data | x | 7-8 |
|  |  | - Statistical methods used for additional analyses, such as subgroup analyses and adjusted analysis | x | 7-8 |
|  |  | - Methods for imputing missing data, if used | N/A |  |
|  |  | - Statistical software or programs used | x | 8 |
| **RESULTS** | | |  |  |
| Participant flow | 12 | - Flow of participants through each stage of the study: enrollment, assignment, allocation and intervention exposure, follow-up, analysis (a diagram is strongly recommended) | x | 34 |
|  |  | - Enrollment: the numbers of participants screened for eligibility, found to be eligible or not eligible, declined to be enrolled, and enrolled in the study | x | Supp Fig 1 |
|  |  | - Assignment: the numbers of participants assigned to a study condition | x | Supp Fig 1 |
|  |  | - Allocation and intervention exposure: the number of participants assigned to each study condition and the number of participants who received each intervention | x | Supp Fig 1 |
|  |  | - Follow-up: the number of participants who completed the follow-up or did not complete the follow-up (i.e., lost to follow-up), by study condition | x | Supp Fig 1 |
|  |  | - Analysis: the number of participants included in or excluded from the main analysis, by study condition | x | Supp Fig 1 |
|  |  | - Description of protocol deviations from study as planned, along with reasons | N/A |  |
| Recruitment | 13 | - Dates defining the periods of recruitment and follow-up | x | 7 |
| Baseline data | 14 | - Baseline demographic and clinical characteristics of participants in each study condition | x | Table 1 |
|  |  | - Baseline characteristics for each study condition relevant to specific disease prevention research | x | Table 1 |
|  |  | - Baseline comparisons of those lost to follow-up and those retained, overall and by study condition |  | Not available |
|  |  | - Comparison between study population at baseline and target population of interest |  | Not available |
| Baseline equivalence | 15 | - Data on study group equivalence at baseline and statistical methods used to control for baseline differences | x | 6 |
| Numbers analyzed | 16 | - Number of participants (denominator) included in each analysis for each study condition, particularly when the denominators change for different outcomes; statement of the results in absolute numbers when feasible | x | 8 |
|  |  | - Indication of whether the analysis strategy was “intention to treat” or, if not, description of how non-compliers were treated in the analyses | x | 6 |
| Outcomes and estimation | 17 | - For each primary and secondary outcome, a summary of results for each estimation study condition, and the estimated effect size and a confidence interval to indicate the precision | x | 8-9 |
|  |  | - Inclusion of null and negative findings | x | 8-9 |
|  |  | - Inclusion of results from testing pre-specified causal pathways through which the intervention was intended to operate, if any | N/A |  |
| Ancillary analyses | 18 | - Summary of other analyses performed, including subgroup or restricted analyses, indicating which are pre-specified or exploratory | x | 8-9 |
| Adverse events | 19 | - Summary of all important adverse events or unintended effects in each study condition (including summary measures, effect size estimates, and confidence intervals) | N/A |  |
| **DISCUSSION** | | |  |  |
| Interpretation | 20 | - Interpretation of the results, taking into account study hypotheses, sources of potential bias, imprecision of measures, multiplicative analyses, and other limitations or weaknesses of the study | x | 9-10 |
|  |  | - Discussion of results taking into account the mechanism by which the intervention was intended to work (causal pathways) or alternative mechanisms or explanations | x | 9-10 |
|  |  | - Discussion of the success of and barriers to implementing the intervention, fidelity of implementation | x | 9-10 |
|  |  | - Discussion of research, programmatic, or policy implications | x | 9-11 |
| Generalizability | 21 | - Generalizability (external validity) of the trial findings, taking into account the study population, the characteristics of the intervention, length of follow-up, incentives, compliance rates, specific sites/settings involved in the study, and other contextual issues | x | 11 |
| Overall evidence | 22 | - General interpretation of the results in the context of current evidence and current theory | x | 12 |

# = Number; N/A = Not applicable.

#### Supplemental Table 2. Percentage of patients with documented SIC in each primary care clinic (N = 13), by study group and time period

| **Study**  **group** | **Primary care clinic**  ID^a^ (N^b^) | **SES^c^** | **SIC training approach** | | **Other SIC training** | **Period 1** | | | **Period 2** | | **Period 3** | | |
| --- | --- | --- | --- | --- | --- | --- | --- | --- | --- | --- | --- | --- | --- |
|  |  |  |  |  |  | **patients with at least one visit**  *n* | | **patients with documented SIC**  *n* (%) | **patients with at least one visit**  *n* | **patients with documented SIC**  *n* (%) | **patients with at least one visit**  *n* | | **patients with documented SIC**  *n* (%) |
| **Intervention** | A  (N = 270) | Privileged | Team-based^d^ | | No | | 221 | 11 (5.0) | 158 | 9 (5.7) | 222 | 9 (4.1) | |
|  | B  (N = 276) | Privileged | Clinician-focus^e^ | | No | | 216 | 3 (1.4) | 160 | 9 (5.6) | 246 | 12 (4.9) | |
|  | C  (N = 188) | Under-privileged | Team-based | | No | | 163 | 6 (3.7) | 110 | 4 (3.6) | 130 | 7 (5.4) | |
|  | D  (N = 354) | Under-privileged | Clinician-focus | | No | | 290 | 9 (3.1) | 197 | 5 (2.5) | 305 | 17 (5.6) | |
|  | E  (N = 254) | Under-privileged | Team-based | | No | | 201 | 8 (4.0) | 145 | 6 (4.1) | 235 | 10 (4.3) | |
|  |  | | | **Total (*n* or %)** | | | 1091 | 37 (3.4) | 770 | 33 (4.3) | 1138 | 55 (4.8) | |
| **Control** | F  (N = 87) | Privileged | N/A | | No | | 63 | 0 (0) | 43 | 1 (2.3) | 78 | 0 (0) | |
|  | G  (N = 81) | Privileged | N/A | | No | | 66 | 2 (3.0) | 50 | 4 (8.0) | 70 | 2 (2.9) | |
|  | H  (N = 87) | Privileged | N/A | | Yes | | 54 | 1 (1.9) | 44 | 0 (0) | 76 | 3 (3.9) | |
|  | I  (N = 222) | Under-privileged | N/A | | No | | 172 | 10 (5.8) | 112 | 6 (5.4) | 192 | 8 (4.2) | |
|  | J  (N = 139) | Under-privileged | N/A | | No | | 116 | 1 (0.9) | 92 | 11 (12.0) | 123 | 25 (20.3) | |
|  | K  (N = 83) | Under-privileged | N/A | | No | | 62 | 2 (3.2) | 47 | 8 (17.0) | 66 | 3 (4.6) | |
|  | L  (N = 246) | Under-privileged | N/A | | Yes | | 162 | 9 (5.6) | 118 | 21 (17.8) | 200 | 14 (7.0) | |
|  | M  (N = 81) | Under-privileged | N/A | | No | | 52 | 0 (0.0) | 45 | 0 (0) | 60 | 0 (0) | |
|  |  | | | **Total (*n* or %)** | | | 747 | 25 (3.3) | 551 | 51 (9.3) | 865 | 55 (6.4) | |

SES = socioeconomic status based on the Material and Social Deprivation Index: "privileged clinics” were situated in geographical catchment areas determined by their postal codes with an MSDI ranging from 1 to 3, and “under-priviledged” clinics were situated in geographical catchment areas with an MSDI of 4 or 5; ID = identification; SIC = serious illness conversation; N or *n* = number; % = percentage; N/A = not applicable.

NOTE:

^a^ID letter assigned to each primary care clinic.

^b^Total number of eligible patient records reviewed at the primary care clinic.

^c^Socioeconomic status of each primary care clinic was determined based on the area-based Material and Social Deprivation Index (MSDI) for public health in Quebec, which is compiled by the Bureau d'information et d'études en santé des populations of the Institut national de santé publique du Québec (INSPQ) using data from the 2021 Canadian census data (publicly available at <https://www.inspq.qc.ca/en/deprivation/material-and-social-deprivation-index>).

^d^The team-based approach (or model) was developed using the Shared Decision Making Model as a framework to adapt the Ariadne Labs’ original SIC training and engage interprofessional primary care teams in SICs.

^e^The clinician-focus approach consists of the Ariadne Labs’ original SIC training ([www.ariadnelabs.org](http://www.ariadnelabs.org)).

#### Supplemental Figure 1. Study flow diagram

Assessed for eligibility (*n* = 3,740)

Excluded (*n* = 1,372)

Not meeting inclusion criteria

Analyzed (*n* = 1,342)

Excluded from analysis (*n* = 0)

Allocated to intervention (*n* = 1,342)

Allocated to control (*n* = 1,026)

Analyzed (*n* = 1,026)

Excluded from analysis (*n* = 0)

**Allocation**

**Analysis**

Included (*n* = 2,368)

**Enrollment**

Patients aged ≥ 65 years old with at least one clinical visit during the study periods^a^ (*n* = 45,008)

NOTE.

^a^Period 1: The six months preceding the onset of the COVID-19 pandemic (September 12, 2019 to March 12, 2020); Period 2: From the start of the pandemic until the release and province-wide email distribution of the basic SIC toolkit by the Quebec Ministry of Health (March 13, 2020 to April 24, 2020); and Period 3: The six months following dissemination of the SIC toolkit (April 25, 2020 to October 25, 2020).

#### Additional information regarding the SIC training program

The SIC training program includes a structured SIC guide, patient identification strategies, training and coaching interventions for HPs, standardized SIC documentation, implementation support, and documentation assistance for families and HPs.^1,2^ Initially designed for the outpatient oncology setting the SIC program has been adapted for multiple settings,^3^ including primary care. Ariadne Lab’s SIC program has also been adapted for and implemented within interprofessional primary care teams,^4,5^ and has been found to be well accepted by care team members and effective in improving their likelihood of engaging in ACP.^6^

Both the intervention and control groups had access to the basic online SIC toolkit, as it was disseminated by the Ministry of Health across all primary care clinics at the start of the COVID-19 pandemic (publicly available since April 24, 2020; <https://publications.msss.gouv.qc.ca/msss/document-002933/>). The toolkit contained open access to educational materials, which included recommendations on how to identify vulnerable patients at high-risk of mortality, clinical guidelines on how to assess and determine levels of care, a short version of the SIC Guide adapted from Ariadne Labs’ SIC program, and information linking HPs to other available training resources. No additional training was provided to accompany the toolkit to either group.
